# Access to autism services and support needs in Morocco: Perspectives of autistic adults, caregivers, service providers, and civil society

**DOI:** 10.1101/2025.03.28.25324760

**Authors:** Oumnia Bouaddi, Afaf Affane, Chaimae Elmalki, Yassine Souidi, Mohamed Khalis

## Abstract

**Background:** In Morocco, limited research has explored access to ASD care. This study aims to examine access to ASD diagnosis, interventions, and support in Morocco, identify barriers and unmet needs, and provide grassroots recommendations for policy and practice.

**Methods:** A qualitative study using in-depth interviews was conducted between October 2024 and March 2025 in three Moroccan regions. Participants included caregivers, healthcare professionals, and civil society actors involved in ASD care. Data were collected in Moroccan Darija, transcribed, and analyzed thematically to identify key themes.

**Results:** A total of 37 participants were included (8 autistic adults, 11 caregivers, 6 civil society representatives, and 15 healthcare professionals). Participants reported delays in ASD diagnosis, attributed to poor awareness and centralized specialist services. Parents reported financial hardships and stigma, including rejection within families and communities, which exacerbated caregiving burdens. Public healthcare services were described as limited, with long waiting lists and inadequate training for professionals, prompting many families to seek costly private care. Civil society organizations reportedly provided essential support, but faced funding constraints and limited reach. Educational integration was deemed limited by untrained staff, insufficient resources, and high costs and non-availability of educational assistants. Services for adults with ASD were severely lacking with limited vocational and profesisonal opportunities.

Conclusions

This study revealed mutliple financial, logistical, and societal barriers limiting access to ASD care across the lifecourse. There is need for increased public commitment and funding, decentralized and inclusive services, capacity-building initiatives, and improved educational and professional integration.

## 1. Introduction

Autism spectrum disorder (ASD) is a complex and poorly understood neurodevelopmental disorders with a multifactorial etiology involving genetic, neurobiological, environmental, neurodevelopmental, and psycho-affective components (*Autism Spectrum Disorder - National Institute of Mental Health (NIMH)*, n.d.). It is characterized by impairments in social interactions, communication, and the presence of repetitive stereotyped behaviors. In addition to these symptoms, other conditions may co-occur, - such as epilepsy, intellectual disability, sensory deficits, motor disorders, and psychiatric conditions, particularly anxiety, depression, attention deficit hyperactivity disorder (ADHD) (*Autism Spectrum Disorder - National Institute of Mental Health (NIMH)*, n.d.). Autism affects all social classes, with a marked predominance in boys compared to girls (Zeidan et al., 2022a). Currently, there is no treatment that directly targets the causes of autism. However, medications are used to manage certain associated symptoms, in association with educational therapies. Global and regional estimates of ASD prevalence are scarce. In 2023, a global systematic review and meta-analysis across 59 studies pooling existing literature on ASD (including Asperger’s and previous classifications) found that the pooled global prevalence between 1994 and 2019 was 0.72% (95% CI = 0.61– 0.85)(Talantseva et al., 2023). Previous literature also highlights an increase in prevalence estimates over time and higher rates in the Western world (Talantseva et al., 2023; Zeidan et al., 2022b). This reported difference has been attributed to an inability to diagnose ASD due to a shortage of available diagnostic services in low and middle income settings, rather than a natural worldwide variation in the incidence of ASD in its different forms (Samadi, 2022a). ASD has significant negative consequences not only for the affected individuals but also for caregivers (Turnage & Conner, 2022). Caregivers face a particularly heavy burden, because addressing the diverse needs of an autistic person often requires a multidisciplinary approach (van Niekerk et al., 2023). Despite the health and social consequences associated with ASD, it remains an understudied area of research, particularly from social and care perspectives, and especially in low-and middle-income countries (LMICs). These regions often have different social and care structures and are more likely to experience chronic health system challenges, such as shortages of allied health professionals and widespread societal stigma, which further contribute to delays in ASD diagnosis and care (Samadi, 2022a).

In Morocco, ASD is a disorder with limited epidemiological research. A few small-scale studies have attempted to describe the profiles of patients or the needs of caregivers. For instance, one study involving 90 parents of children with ASD found that 61% of the children were boys, with a sex ratio of 2.6 (Sefrioui et al., 2023). Early signs of autism appeared at the age of 18 months for 22% of the sample (Sefrioui et al., 2023). Similarly, another study conducted with 131 Moroccan caregivers of autistic children across five rural and urban regions identified several caregiver needs, including the improvement of healthcare and educational services for autistic children and training programs to develop their social, communication, intellectual, and autonomy skills (de Jonge et al., 2024). This study also highlighted persistent challenges in accessing services due to their unavailability, long waiting lists, lack of information, and high costs (de Jonge et al., 2024). These studies remain relatively limited and do not adequately capture the experiences of parents and caregivers, nor those of healthcare professionals (psychologists, child psychiatrists, etc.) who are closely involved in autism care. The primary objective of this study is to understand access to diagnosis, interventions, and support for individuals with ASD in Morocco, as well as to identify the challenges faced and the unmet needs of individuals with ASD and their families, through the lens of austic adults, caregivers, healthcare professionals, and civil society actors involved in providing care. A secondary objective is to identify grassroots recommendations from these stakeholders. Therefore, the findings of this study hold significant and immediate policy and practice implications for policymakers, and will contribute to the development of the ongoing policy.

## 2. Methods

### Study design and setting

This is a phenomenological qualitative study conducted in three regions in Morocco; Tanger-Tétouan-AlHoceima, Marrakech-Safi and Rabat-Salé-Kénitra, between October 28, 2024, and March 16, 2025.

### Study population and recruitment of participants

We used purposive sampling for this study. The study population included: (i) adult individuals living with ASD (ii) adult parents and caregivers of individuals (children and adults) living in the three study sites, (iii) healthcare professionals involved in caring for autistic children and adults, and (iv) civil society actors and elders working in organizations that provide care for autistic children, adults, and their families. Parents, caregivers, and civil society actors were recruited through the national civil society organization *Collectif Autisme Maroc*. *Collectif Autisme Maroc* is a national network of parents’ associations whose mission is to promote the rights of people with autism. Healthcare professionals were recruited purposively and through snowball sampling using the research team’s established network in tertiary healthcare facilities in the three regions. We included a diverse range of professionals, including psychomotor therapists, speech and language therapists, child psychiatrists, and psychologists due to the multidisciplinary nature of autism care.

### Data collection

In-depth interviews were conducted with caregivers, health providers and civil society actors whereas focus group discussions were done with autistic adults. All sessions were carried out in Moroccan Darija (dialect) by members of the research team (OB and CE) via telephone or online via videoconference. Four topic guides were developed for the study subpopulations. These guides were created by two members of the research team (OB) and validated by MK. The topic guides were informed by existing literature on autism services and included questions exploring perceptions of autism services from pre-diagnosis to post-diagnosis and from childhood to adulthood. The guides aimed to identify gaps or barriers in service access and coordination. Questions were designed to capture both the experiences and challenges faced by caregivers and autistic adults across the life course, including access to services beyond healthcare (e.g., schooling, recreational activities) and support services for parents themselves. We also collected socio-demographic data, including age, gender, region, level of education, and employment status for caregivers and autistic adults, and age, gender, region, and specialty for healthcare professionals. Each interview lasted between 30 and 45 minutes, during which field notes were taken. For autistic adults, the topic guide was shared one week prior to the session to ensure predictability and familiarity with the questions by participants.

### Data analysis

Data analysis was performed concurrently with data collection, which allowed for the iterative development of key themes. We used a flexible and rapid qualitative thematic analysis approach (Vindrola-Padros, 2021). Field notes were reviewed multiple times and coded manually by OB. Codes were grouped into broader concepts of barriers and challenges across the life course (pre-diagnosis, post-diagnosis, adulthood). Themes were developed from thematically significant codes. Initial codes were revisited and reorganized as more data was collected to refine the findings.

### Ethical considerations

The study protocol was approved by the Ethics Committee of the Mohammed VI University of Sciences and Health (CE/UM6SS/63/24). Verbal and informed consent was obtained from all participants before the start of each interview. No identifiable data was collected. Participants were reminded that their participation was voluntary and that they could withdraw from the study at any time.

## 3. Results

### Socio-demographics characteristics

We conducted 11 semi-structured interviews with parents of children and adults with ASD, 6 representatives from civil society organizations, and 15 professionals from various backgrounds (Table 1). We also conducted one FGD with eight autistic adults. Among the parents interviewed, 6 out of 11 were men, with an average age of 50.1 ± 8.21 years. The average age of their children/adults with ASD (autism spectrum disorder) at the time of the interviews was 17.67 ± 6.77 years. The majority of participants had higher education (9/11) and were employed or retired (10/11). The average age of the healthcare professionals was 33.1 ± 6.2, and 13/15 were female. A diverse range of professionals was included, including 4/15 speech therapists, 3/15 psychomotor therapists, and 3/15 psychologists. For autistic adults, the mean age was 23.8 ± 6.09 years. Six out of eight participants were males, five were from the Tanger--Al Hoceima region, and four were pursuing higher education or vocational training, while the remaining participants had dropped out of primary school or high school.

**Table 1.** General characteristics of participants.

| Variables | Parents (N=11) |
| --- | --- |
| **Parents (N=11)** |  |
| **Age (mean ± SD)** | 50.1 ± 8.21 |
| **Age (children) (mean ± SD)** | 17.67 ± 6.77 |
| **Sex** |  |
| Male | 6 (54.5) |
| Female | 5 (45.5) |
| **Level of education** |  |
| None | 1 (9.1) |
| Higher education | 9 (81.8) |
| Not disclosed | 1 (9.1) |
| **Employment status** |  |
| Employed/Retired | 10 (90.9) |
| Unemployed | 1 (9.1) |
| **Region** |  |
| Rabat-Salé-Kénitra | 3 (27.3) |
| Tanger-Tétouan-Al Hoceima | 5 (45.5) |
| -Safi | 3 (27.3) |
| **Health professionals (N=15)** |  |
| **Age (mean ± SD)** | 33.1 ± 6.2 |
| **Sex** |  |
| Female | 13 (86.7%) |
| Male | 2 (13.3%) |
| **Region** |  |
| Rabat-Salé-Kénitra | 5 (33.3%) |
| Tanger-Tétouan-Al Hoceima | 5 (33.3%) |
| Marrakech-Safi | 5 (33.3%) |
| **Profession** |  |
| Speech therapist | 4 (26.7%) |
| Psychomotor therapist | 3 (20.0%) |
| Psychologist | 3 (20.0%) |
| Child psychiatrist | 2 (13.3%) |
| Neurologist | 1 (6.7%) |
| Social worker | 1 (6.7%) |
| Mental health nurse | 1 (6.7%) |

Four major themes were identified (Figure 1) which are described below.

**Figure 1.**
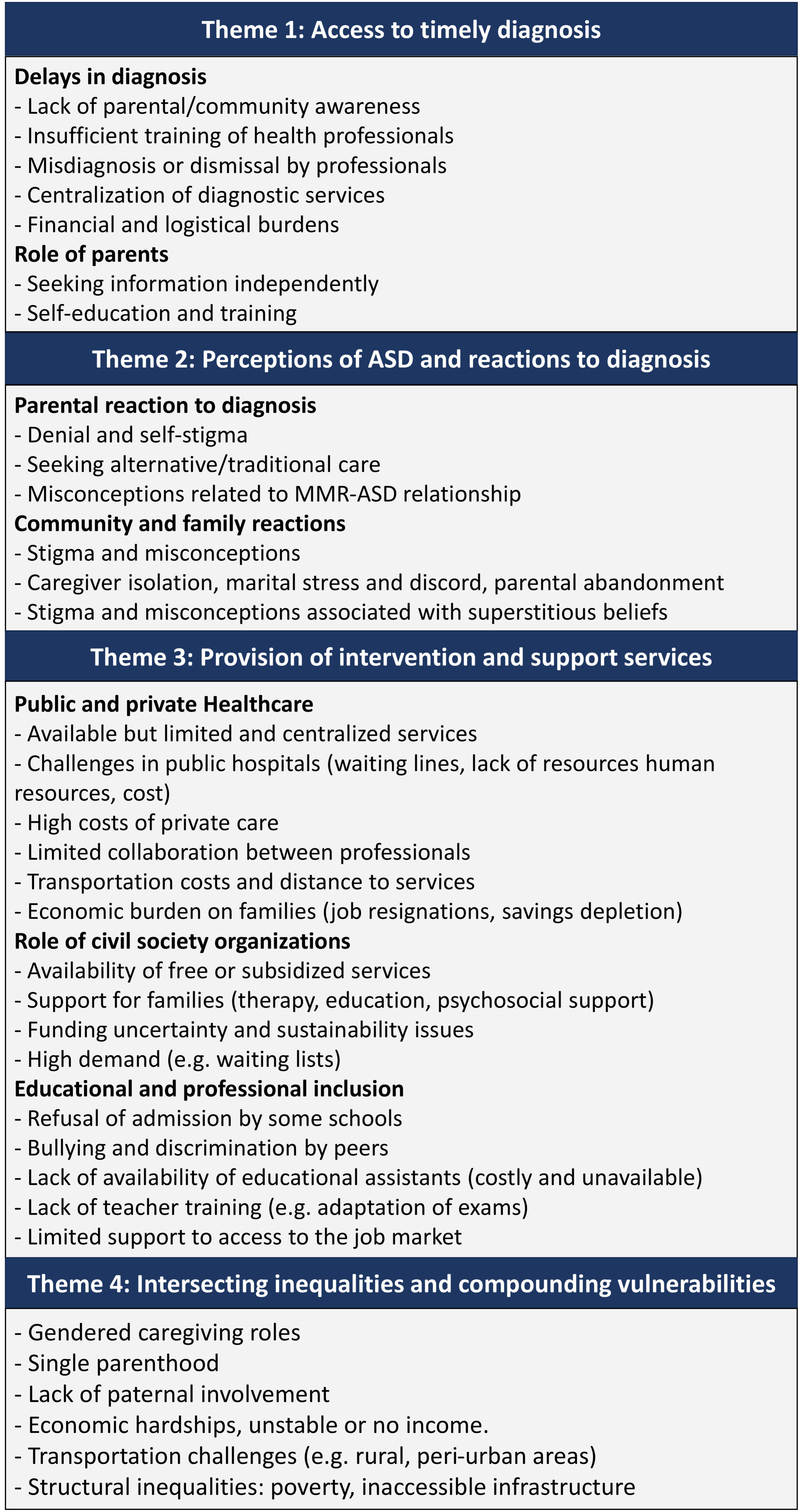
Main themes and codes related to access to diagnostic, intervention and support services by individuals with ASD and their families

### Theme 1: Access to timely diagnosis

While recounting their experiences from the onset of symptoms to diagnosis and the current situation of their children, parents and ausxtic adults described several barriers they encountered in seeking a diagnosis for their children. Many parents reported seeking diagnostic services at a time when autism was “less known,” which led to significant delays in obtaining a diagnosis, ranging from 1 year to more than 5 years across participants. Poor parental awareness and limited community knowledge about ASD were also mentioned as contributing factors to the delays in diagnosis:

*“At 8 months, I first noticed something wasn’t normal, but everyone said there was no problem.”– Parent*

*“There are parents who say, ‘He looks after his father, he will start talking eventually, it’s just a delay.’” – Nurse specializing in mental health,*

Participants reported several other factors contributing to delayed diagnosis including poor information about available services, uncertainty about where to go, and a lack of knowledge, awareness, and training among healthcare professionals in diagnosing ASD. To address the lack of information about ASD, several parents mentioned seeking information themselves, paying for online training or pursuing formal education to better understand their child’s condition as well as seeking a diagnosis abroad. For several autistic adults and caregivers, the first point of contact with healthcare professionals—often pediatricians, or psychiatrists—did not result in a diagnosis, resulted in a misdiagnosis, or was dismissed as normal developmental delays in verbal communication:

*“It [getting a diagnosis] was not easy. It was very difficult for several reasons, lack of doctors who are aware of and specialists in ASD… the lack of services for ASD, such as in my hometown.” – Autistic adult*

*“We took him to the pediatrician, and she told us it’s normal—just a normal delay.” – Parent, Marrakech*

*“Unfortunately, even our family pediatrician didn’t notice anything. He didn’t know. He just focused on growth and vital signs. We started searching for answers abroad…they finally gave us a proper diagnosis.” – Parent,*

In most cases, an eventual diagnosis was made in urban centers by few specialists renowned for their expertise in ASD, due to centralizeation of trained health professionals:

*“My parents noticed that I was different and took me to a pediatrician, but sadly, he was unable to diagnose me. He then suggested that my parents go to Rabat or Casablanca, but at that time, ASD was not well-known. Due to the cost and difficulty of transportation, we didn’t go immediately.” – Autistic adult*

*“We took him to the pediatrician. He said it was nothing, just delayed speech. I started comparing him to his brother’s development and noticed a delay. Then, I took him to Rabat to see a professor there.” – Parent*

Even before the start of intervention services, autistic adults and parents reported that the journey to obtain a diagnosis was financially burdensome. In extreme cases, as reported by one participant, families relocated to Rabat for months to seek a diagnosis by experienced professionals:

*“I went to the pediatrician, and he said my son has autism. I asked what that meant, and he said he would refer me to a psychiatrist. I went to her, but she didn’t give me clarity. I then went to Rabat, where I stayed for a while, moving between doctors. I traveled Rabat very well for my son, and eventually, we confirmed he truly has autism.” – Parent*

Health professionals noted that while there were significant delays in diagnosis in the past, they are now seeing more younger cases, which they attributed to increased awareness among parents. They also emphasized that delayed diagnosis remains a major challenge, as it undermines the effectiveness of care and negatively impacts prognosis:

*“More people are presenting with ASD, and the diagnosis is being made earlier now. We are seeing more children.” – Child psychiatrist*

### Theme 2: Perceptions of ASD, reactions to diagnosis and experiences of stigma

While participants did not explicitly affirm a belief in the MMR vaccine as a cause of autism, it was mentioned as a potential factor considered. Some parents associated the appearance of symptoms with the timing of vaccination:

*“The mother thought that it may be because of the MMR vaccine” – Parent*

*“After the vaccine of 9 months [MMR], he changed, I noticed is by his stare, his eyes have sort of died, he lost visual fixation, suddenly, started entering a state of isolation, and having crises” – Parent*

Once the diagnosis was established, several parents mentioned the difficulty of accepting the long-term nature and untreatability of ASD. This was often accompanied by denial, self-stigma, and the pursuit of alternative care methods. For example, one mother mentioned seeking traditional services:

*“The majority of parents are in denial, we have a lot of problems” – Speech and language therapist*

*“Now I tell the mothers do not make the same mistakes I did, I was not listening, I was following Foqha and Salihin” – Parent*

Civil society actors and some parents described lived expeiences of how a confirmed ASD diagnosis was met with rejection by the father, abandonment and refusal to take responsibility for the child, leaving the caregiving burden on the mother or divorcing her to escape responsibility:

*“In the centre, we have cases whose parents have abandoned them, it hurts me so much, they say, he is not my son, the father neglects a human obligation, drops all responsibility on the mother, we have so many cases like these.” – Parent and civil society actor*

The challenge in accepting the diagnosis was often exacerbated by stigma and societal misconceptions, and superstitious beliefs within households, neighborhoods, and the broader community. For instance, one mother, who also works as a civil society advocate, shared examples of parents of autistic children being forced to relocate every few months due to neighbors persistently complaining to landlords about the noise. In many cases, ASD diagnosis reportedly led to marital discord and even divorce:

*“Lack of awareness in society, we say he is hit by a spirit, the family starts believing that they have been cursed, now there is a bit more awareness” – Parent*

Autistic adults shared that their experiences with bullying, stigma, and discrimination have been ongoing since childhood, both in school and in the broader community. Two participants described how talking to oneself, particularly in public, is often perceived as being’hit by a spirit,’ and they make efforts to use this behavior to help themselves. They also believe that it’s a sign of intelligence. These negative experiences are often worsened by a general lack of understanding of ASD in society, which leads to further social isolation and mental health challenges. Despite the growing awareness of autism in Morocco, participants reported that the social barriers that autistic individuals face persist, due to expectations to conform to norms that do not align with their neurodivergence. Autistic adults emphasized the need to recognize their neurodiversity as opposed to viewing it as a disorder.

*’The challenges I face now include a lack of awareness about ASD. I face bullying and discrimination at school and sometimes in life outside of school. I get called crazy, disabled, and stupid every time I try to play with them. The teachers also say, “You’re a failure, you’re stupid,” and that I move a lot. Another problem is talking to myself. It’s not being crazy; it’s a sign of intelligence, but I try to control it and use it in ways that help me.’ – Autistic adult*

### Theme 3: Provision of intervention and support services in public, private and civil society sectors

#### Public and private healthcare services

At the beginning of their journey to seeking services, parents reported initially turning to public hospitals. However, parents described multiple challenges including long waiting lines, a severe lack of healthcare human resources (such as psychomotor and speech therapists), poor training and, in some cases, stigma and poor interpersonal communication with health professionals. As a result, participants often turned to the private sector, where services were more readily accessible. However, the costs of the later were exorbitant, and the quality of services was reported as suboptimal due to lack of training of private sector health professionals in ASD:

*“ And when you go inside, it’s just 15 minutes. It’s not enough. And when you reach a number of sessions, they tell you,’That’s it.’– Parent*

*“But when you go to the provincial hospital, there is a psychomotrician and a speech therapist, but there’s a long waiting line, and only one speech therapist.” – Parent*

*“There were many challenges: lack of training, even for specialists.” – Parent*

These challenges were also highlighted by health professionals who explained that, in ambulatory care, only a limited number of sessions for speech and psychomotor therapy are offered. However in-hospital specifically for ASD exist, and they offer speech therapy, behavioral therapy, and psychomotor therapy are provided for up to three months, and were perceived as effective work environments.

*“[We provide] only 15 sessions; otherwise, they can pay for the day hospital, which offers up to three months. Most patients we receive have full insurance [AMO Tadamoun]”– Psychomotrician*

Health professionals reported facing challenges in delivering care, including insufficient numbers of professionals and parents interrupting care. This interruption was described as occurring either due to low economic status and inability to pay or, in some cases, due to poor awareness. Additionally, they faced challenges in collaboration, which was described as poor outside of the day hospital setting:

*“The primary challenge, in my opinion, lies in the lack of cooperation from some families, which is often the main reason for abandoning car [..] The professionals involved in the care of autistic children are insufficient in number, which leads to significant delays in the care of many patients.”* – *Psychologist*

*“Unmet needs include financial support, as some families have had to interrupt follow-up care due to economic difficulties.”* – *Psychomotor therapist*

### Service provision by civil society organizations

Autistic adults and caregivers mentioned receiving valuable support from CSOs which offered diverse and tailored health and educational services. While some provided these services free of charge due to public funding from entities such as the Ministry of Solidarity or the National Initiative for Human Development (INDH), other associations faced significant challenges due to a lack of sustainable funding. These associations often relied on beneficiary contributions to cover the costs of human resources, making their services inaccessible to many families. In some centers offering free of charge services (due to public funding) and managed by CSOs, autistic adults and caregivers reported benefiting from high-quality and comprehensive support, including speech and language therapy, psychomotor therapy, sensory integration, individual monitoring, and follow-ups with trained educators. Parallel activities, such as sports and artistic programs, were also available:

*“I received many services from [name of CSO], including speech therapy and psychomotor therapy. It helped me improve my communication skills, integrate into school, and express myself.” – Autistic adult*

*“As I see, the educators in these centers are good. They watch the kids. It makes you feel relieved as a mother, unlike other centers where there is neglect and sometimes violence and exploitation*.

*Those who have money can benefit, and those who don’t, their kids are left out.”– Parent*

*“The association offers a lot of services: sports, speech and language therapy, psychomotor therapy, and education.” – Parent*

However, these free services were mainly reported in urban centers. Waiting lists and increasing demand were a major challenge, and CSO representatives frequently expressed uncertainty about future funding. Many CSO actors voiced concerns that funding gaps could leave families without any support:

*“There is a big waiting list. I spent five years trying to start benefiting. I quit my job so I could be eligible. I risked my future.” – Parent*

*“Our work is limited without continuous institutional support. If the Ministry of Solidarity’s programs stop, it will create problems for us, especially with our human resources.” – Parent*

### Support services for parents and caregivers

Parents reported benefiting from various support services provided by CSOs. These included workshops of parental guidance on ASD, opportunities to observe educators working with their children, and publicly subsidized training programs like *Rafiq* for caregivers and professionals involved in providng care for individuals with ASD. Participants appreciated the applied learning component, where supervisors monitored and guided their practice. However, the program had limited admission slots. Additionally, some parents benefited from individual or group psychosocial support sessions with other parents:

*“I participated in a program called Rafiq, organized by the Ministry of Solidarity. They trained us on how to accompany our child.” – Parent*

*“[We provide] parental guidance. Every specialist does it. Accompaniment is very important. We also train our colleagues. Those trained as part of Rafiq do this for the benefit of parents, to explain ASD and ABA.” – Psychomotrician*

### Logistical and financial challenges

The financial burden of accessing care emerged as a major challenge. Health professionals noted that ambulatory care for children with ASD is provided for a fee. In cases where patients do not benefit from an insurance scheme that covers the cost, they are required to prepay and then seek reimbursement, which health professionals perceived as a significant barrier, leading some families to drop out of care.

*“For AMO Solidaire [health insurance scheme for the most disadvantaged], the day hospital is free, but for other health insurance schemes you have to pay upfront and then they reimburse some of it. This is a big problem, as some parents do not have the means. Even if the waiting list reaches them and it’s their turn to benefit, they cannot..” – Speech and language therapist*

As most parents in the study had given up on public healthcare or had already utilized the allocated number of sessions, the high costs expressed by parents were mainly associated with private care or services provided by fee-based associations. Even middle-class families and highly educated parents reported depleting their savings to cover these expenses. Despite the high cost of care, many parents, particularly mothers, had to leave their jobs to take on full-time caregiving responsibilities for their children, adding another layer of financial hardsip:

*“Seventeen years of work—we sacrificed financially and emotionally. Autism impoverishes you.” – Parent*

*“We [parents] make exceptional efforts, more than educational assistants.. I wish they were trained. I quit my job to be an educational assistant for my child.” – Parent*

*“The mother [my wife] resigned to look after her [our child].” – Parent*

With some publicly funded, free-of-charge centers now available, most parents sought to take their children to these facilities despite limited access due to long waiting lists. Transportation expenses also posed a significant barrier for accessing both public health facilities and ASD centers, particularly because of the centralization of services in urban areas. For example, one participant said that an entire province lacked a single child psychiatrist, while a health professional mentioned that a whole region had only one child psychiatry department:

*“First, there is a problem with transport. I stay there all day. Transport is the main issue. I used to take her to the swimming pool, but I stopped because of the distance. There are things I sacrifice—I don’t do for myself so I can provide for my child.” – Parent*

*“In the whole province of Essaouira, there is no child psychiatrist. Marrakesh, 175 km away, is the nearest city to see a child psychiatrist, and it takes three hours to get there.” – Parent*

*“There is only one child psychiatry department in the entire region—the only service for child psychiatry is in Rabat. It’s very centralized.” – Psychomotrician*

Both parents and CSO representatives identified the lack of trained professionals as a significant barrier. Parents noted that services provided by free of charge publicly supported centers managed by CSOs offered higher-quality services due to the well-trained allied health professionals specializing in ASD. CSOs reported facing challenges in recruiting trained educators, psychologists, and other professionals. Consequently, many associations invested in capacity-building initiatives to raise service quality to acceptable standards:

*“The prevalence of ASD has increased a lot. We see many cases, and the waiting list is huge. The problem is that trained professionals are very limited. The association brings people and trains them, but in reality, there aren’t any.” – Parent*

### Access to education across the life-course

Autistic adults, parents and civil society actors shared experiences of school principals, in both public and private sectors, refusing to admit children with ASD despite laws mandating the admission of children with mild cases. Even when children were admitted, schools often required parents to provide an educational assistant, which parents reported as costly and challenging due to the lack of publicly trained and qualified assistants. Some schools also reportedly refused to allow educational assistants to accompany children:

*“I was happy that I was going to school, wearing my clothes and carrying my bag on my back. But they didn’t let me in. I cried a lot until the association intervened, and they finally allowed me to attend with an education assistant.” – Autistic adult*

*“We enrolled her in school, they refused her at first, we started teaching her ourselves” –Parent,*

*“When the child doesn’t sit down, the educational assistant becomes mandatory, but it’s very expensive […] Now, the child has been denied school in another indirect way due to the lack of an educational assistant.” – Parent*

Autistic adults and parents also reported that many teachers lacked training in ASD and ABA, leading to poor understanding of the specific needs of persons with ASD. Instances of mistreatment, lack of engagement in adapting exams, and even bullying by peers, especially in secondary and high schools, were frequently mentioned. Adults described experiences of belittlement and being denied space to express themselves in educational settings. Some parents felt compelled to withdraw their children from schools due to these challenges. Autistic adults and parents also stressed the lack of engagement from schools and educational institutions to adapt exams and assessments despite the presence of directives and guidelines; this was also the case for higher education:

*“When you’re in a group project, someone always tries to control you, push you to the bottom of the pyramid, and make decisions for you. There’s no respect. All doors close in your face, and you fall into depression, feeling like you can’t go on. You feel like everyone is against you. They don’t see your abilities, and sometimes people make fun of you.” – Autistic adult*

*“It [bullying] happened throughout my entire educational journey, but in secondary school, it became extreme and more severe. I used to go to the teacher’s office to talk about the maltreatment, or to the library to read books that could benefit me. I never thought about leaving my studies.” – Autistic adult*

*“You also find that there is no adaptation of exams in higher education. Last year, I was talking to teachers at the institute, and sometimes they told me,’You’re smart, you can pass, we don’t need to do anything’”– Autistic adult*

Parents and civil society actors commended programs put in place by The Ministry of Education has implemented initiatives like integration classes and resource rooms (“*salles de ressources*”) for children with autism, which tailored support and activities, and are increasingly availabile in public schools. However, parents raised concerns about mixing disabilities in these classes and the lack of trained teachers overseeing these classes. In response to this challenge, some associations organize community-based training programs on ASD and ABA for the benefit of teachers overseeing these resource rooms:

*“The problem with the salle des ressources is that they mix all disabilities together.” – Parent Services for youth and adults with ASD*

Parents, autistic adults and health professionals expressed a severe lack of health, education and integration services for adults and quasi-absence of visibility over their future, although autistic adults all stressed their desire and ambition to pursue higher educcation. Only one CSO representative mentioned services for adults, specifically a vocational training center. In other cities, no services were available for adults with autism and there is an absence of educational assistants in higher education. One autistic adult pursuing higher education also expressed frustraion over the lack of adaptation of exams in these settings:

*“There are no services for autistic adults, neither in child psychiatry nor in psychiatry.” – Child psychiatrist*

*“There is also a center for vocational training. My daughter benefits; she is learning esthetics. She wanted to learn journalism. Until now, though, there are no educational assistants to accompany autistic adults in higher education institutions.” – Parent*

Beyond higher education, autistic participants expressed frustration with the absence of services for adults throughout the life course, particularly regarding access to the job market and vocational training that enables them to thrive. Participants placed significant emphasis on their desire for and right to independence and voiced frustration over instances where others undermined that right in work, internship, and study settings, for example, by taking over their tasks or not valuing their skills:

*“For those studying, no training programs are available that prepare you for work. There is a need for better integration into the job market.” **—** Autistic adult*

*“When I was going to intern, I found that people would try to do my work for me. In my experience, they took away tasks that I was capable of doing.” **—** Autistic adult*

### Theme 4: Intersecting inequalities and compounding caregiver vulnerabilities

Discussions with parents and civil society actors revealed how intersecting identities and structural inequalities amplify caregiver vulnerabilities. Factors such as being female, and single parenthood were key contributors to caregiver burden, with mothers disproportionately carrying childcare responsibilities.

*“Neither me nor my husband have an income.” – Parent*

*‘Sometimes, kid go to beg with their children in the streets, this is tragic for us as an association” – Parent and civil society actor*

Despite fathers’ physical presence in many families, a lack of paternal involvement left mothers navigating both caregiving and societal expectations, deepening their marginalization:

*“All the responsibility is on the mother. We need to talk to fathers. As a mother, you watch her, wash her, change her. God give us patience. We don’t go out, and no one invites us anywhere because of these children.” – Parent*

*“Autism is fatherless.” – Parent*

Geographic location added further challenges. Families in rural or peri-urban areas faced limited and costly transportation options, which were often inaccessible for mothers of children with disabilities requiring mobility aids like wheelchairs. Structural barriers, including poverty and a lack of inclusive services—such as accessible roads and public transportation—isolated families and restricted their access to essential support.

### Recommendations to address the needs of children, adults with ASD and their families

Based on the needs expressed by parents and the recommendations suggested by parents, health professionals, community actors, and CSO representatives with lived experience, we have compiled several recommendations for policy and practice to promote access to health and other services for individuals with ASD (see Figure 2).

**Figure 2.**
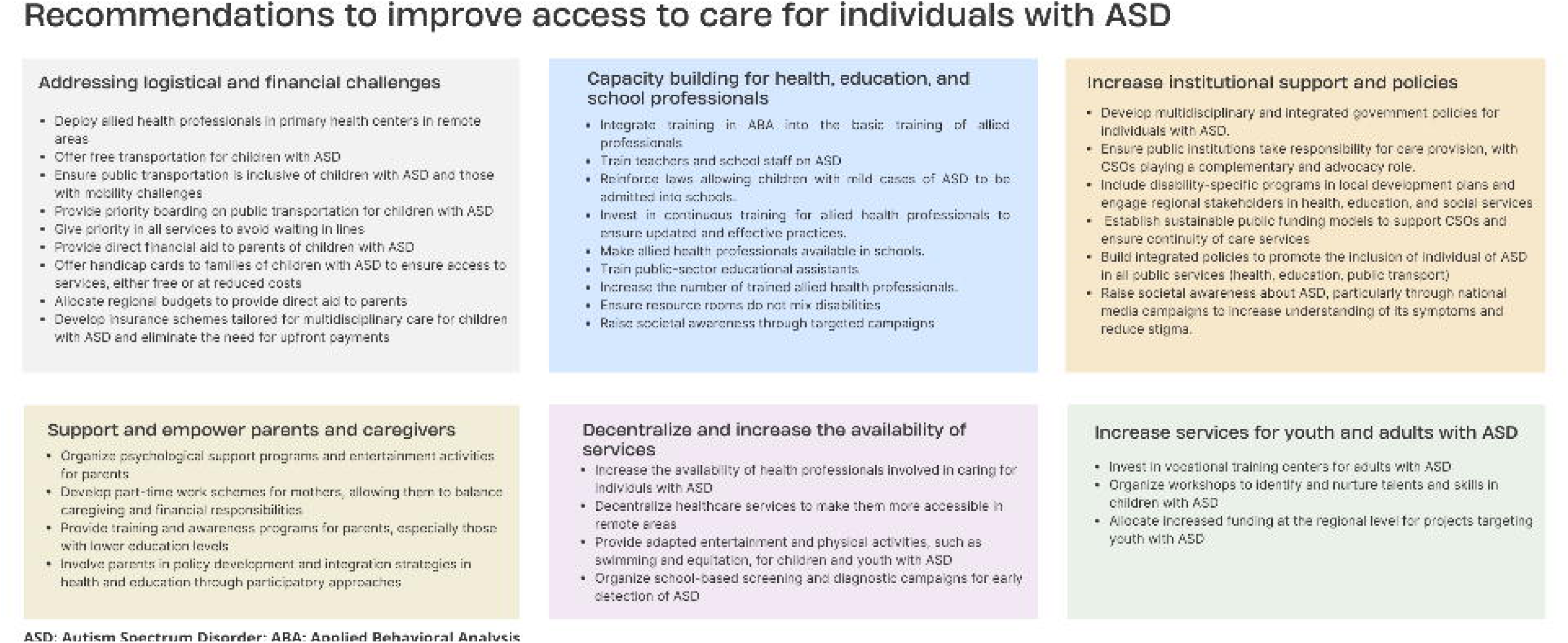
Participant recommendations for policy and practice to promote access to health, education, and other services for individuals with ASD in Morocco

## 4. Discussion

The aim of this qualitative study was to document experiences with ASD diagnostic, intervention and support services, and to understand the challenges encountered in accessing care, including health and education, from the perspective of caregivers, autistic adults, health professionals, and community actors. Participant narratives revealed several challenges experienced across the life course for individuals with ASD, from the onset of symptoms through adulthood. Major delays in diagnosis were described, caused by the heavy centralization of services, a lack of qualified professionals, limited availability of diagnostic services, and poor awareness of the disorder within families and the community. Parents indicated they used a wide range of health and educational services, but findings revealed that service provision in the public health sector was inadequate, insufficient, and often costly for those lacking health insurance and who were required to make upfront payments. Participants reporting from free of charge services by publicly funded CSOs. However, challenges such as long waiting lists and uncertain funding were expressed by CSO actors as major barriers limiting the reach of their services. In addition to health-related challenges, access to education was impacted by poor staff training, lack of qualified educational assistants, bullying, despite promising initiatives introduced by the government. A quasi-absence of health and education services for youth and adults with ASD was reported with no access to the job market and accompaniment for the transition into adulthood. Finally, the study’s recommendations focused on addressing the identified logistical and financial challenges to care, increasing institutional support, and urging the government to take a more active role in service provision. Additional support, both direct (e.g., cash assistance) and indirect (e.g., parental guidance and training), was emphasized to reduce the financial and emotional burden on caregivers.

A key finding in this study is the delay in diagnosis mainly attributed to the lack of proper training for health professionals and the heavy centralization of services, which prevented individuals from obtaining a diagnosis at the first point of contact. This is consistent with studies in LMICs reporting considerable delays between the point of first contact and eventual diagnosis, as well as subsequent early interventions (Samadi, 2022b). Another factor exacerbating delayed diagnosis was cultural beliefs and societal beliefs around autism. Similar findings were decribed in the African continent, showing that such beliefs can lead to delays in seeking medical attention and turning to traditional and alternative care (Aderinto et al., 2023). Cultural beliefs and norms were also found to influence care expectations by caregivers as they may put emphasis on conformity and integration over individual progress and expression (Walsh et al., 2020). To address these beliefs, one key recommendation from this study was to leverage the media to raise awareness about ASD and its symptoms in order to increase demand for timely diagnostic care.

In this study health professionals the implementation of widespread screening campaigns in schools was recommended. In Morocco, there is a quasi-absence of ASD screening campaigns. However, the litterature shows that screening pathways are scarce in both LMICs and high-income settings (Durkin et al., 2015). Screening for autism is expensive, and the literature indicates that gold-standard diagnostic tools used in high-income settings are often too costly for low-income settings (Aderinto et al., 2023; Samadi et al., 2022). As is the case in this study, studies in LMICs report significant delays between the point of first contact and eventual diagnosis, as well as subsequent early interventions (Divan et al., 2021). The literature from LMICs, such as Iraq (Samadi, 2022b) and Chile (Roman-Urrestarazu et al., 2021), shows the potential of short screening instruments that should be validated and could be used to run screening campaigns in Morocco. In addition, leveraging mobile technology and telemedicine, which are gaining significant momentum in Morocco, has great potential to reach a large number of children in need of screening and diagnostic care (Jallal et al., 2023).

The financial and emotional burden, as well as the social cost of care, reported by parents in this study was considerable. This was exacerbated by compounded vulnerabilities, such as being the sole caregiver of the child, living in urban areas far from service facilities, and low economic status. Our findings are corroborated by global litterature showing that caregivers face financial hardships due to the multidisciplinary nature of care and the lifelong need for support, compounded by the lack of appropriate insurance coverage, limited social support, and, in some cases leading to marital problems and separation (Aderinto et al., 2023; Liao et al., 2019). Similarly, several studies in LMICs have reported a high emotional burden on caregivers, particularly mothers, including depression, low quality of life, and anxiety (Ilias et al., 2018; Nuri et al., 2020a, 2020b), often exacerbated by persisting societal and structural stigma (Liao et al., 2019). In the Middle East, a review of studies found that fathers of children with ASD often reported greater financial pressures (Al Khateeb et al., 2019). Globally, financial and emotional stressors related to caring for a child with ASD have been shown to negatively impact participation in social life and contribute to marital problems, as also observed in this study (Divan et al., 2021). In LMICs, the lack of universal health coverage and long-term financial support for services renders autism care prohibitively expensive, thereby limiting access to services and leading to discontinuity in care.

Financial stress has also been linked to long-term emotional distress, as the prolonged nature of care often drives families, even those relatively well-off, into precarious financial situations (Hunt & Watermeyer, 2017). While recent data on funding for ASD diagnosis and care are scarce, a WHO report from 2005 highlighted an overall lack of services focusing on specific disorders (World Health Organization et al., 2005). It found that funding mechanisms in LMICs were unclear; however, a trend of developing specialized services was observed, driven by emerging knowledge and parental advocacy. In Morocco, the situation may be somewhat comparable (World Health Organization et al., 2005). Organized efforts by civil society organizations, combined with increased awareness and evidence-based diagnostic and intervention methods, may have led to improved service availability. However, the availability of services in the public sector remains insufficient, partly due to limited accessibility and the lack of appropriate training for health professionals involved in ASD care. This is consistent with global litterature suggesting low awareness of ASD as a barrier to timely diagnosis and early intervention (Walsh et al., 2020), even among frontline health workers due to limited training opportunities (Shorey et al., 2020; Wang et al., 2020). This calls for investments in continuous education for professionals involved in ASD care and also reforms to integrate applied training in ASD as part of basic training (Divan et al., 2021), as was recommended by health professionals in this study.

The recommendations compiled in this study offer a starting point for improving the availability and quality of care for autism, as well as for increasing the inclusivity of services. While legislation in Morocco grants the right to services for people with disabilities, it was recommended that more active efforts should be made to reinforce this and that it should be supported by real institutional backing and adequate funding. Despite the growing needs of families, the availability and training of health professionals were seen as suboptimal, both in terms of quality and quantity. The involvement of parents as key partners rather than just beneficiaries was recommended in this study and in the litterature on the topic (Divan et al., 2021). In fact, parent-mediated ASD interventions have been tested elsewhere (Rahman et al., 2016; Stahmer et al., 2019) including case studies in the EMRO region which show some promise (Rahman et al., 2016). The peer-to-peer model for guidance, emotional support, building resilience, and navigating services has also shown success. It is not an unfamiliar model in the Moroccan context of ASD care, as parent networks like *Collectif Autisme Maroc* have demonstrated a successful framework that warrants further study. Learning from service delivery and social care experiences in similar contexts is also essential. However, it is important to acknowledge that the biomedical understanding of autism may differ from its societal interpretation, which shapes which models of care are best suited (de Leeuw et al., 2020). Future research should explore this dynamic. Moreover, the role of religious communities in providing emotional support was not explored in this study. Nevertheless, existing literature highlights the potential of faith-based organizations in promoting resilience among caregivers, which presents an area for future exploration.

This is the first qualitative study aiming to understand the landscape of service provision for ASD in Morocco and the first to triangulate perspectives from several stakeholders involved. Despite its strengths, this study has some limitations. The parents recruited for the study were mainly connected through civil society organizations and mostly resided in urban areas, where social networks and resources are more accessible. As a result, the study may not adequately represent parents and caregivers living in remote or rural areas, who may have different levels of access to services. In addition, the study did not include the voices of individuals with ASD themselves, which limits a comprehensive understanding of their lived experiences and needs. Future research should include their perspectives. Finally, the findings of the study may not be directly applicable to other regions in Morocco or to contexts with differing levels of infrastructure, resources, and cultural practices. Expanding the scope of inquiry will ensure more inclusive and culturally relevant recommendations for improving ASD care in Morocco.

## 5. Conclusions

This study highlights that while individuals with ASD in Morocco have access to some services, significant financial, logistical, and societal barriers persist across sectors. Caregivers face a considerable burden, exacerbated by stigma, inadequate support systems, and limited services, particularly for adults with ASD. Autistic adults are faced with limited educational and professional opportunities. Recommendations include increased public funding including direct support to families of children and adults with ASD, decentralized healthcare services, capacity-building initiatives for all professionals interacting with ASD individuals, and improved educational and professional integration across the lifecourse to reduce caregiver burdens. These grassroots findings should be considered in ongoing policy development efforts in Morocco.

## Data Availability

The raw qualitative dataset will not be available. However, additional quotes supporting each theme can be provided upon request from the corresponding author.

## Acknowledgements

We would like to acknowledge all participants who took part for sharing their life experiences with us and actively participating in the study

## Notes

### Competing Interest Statement

The authors have declared no competing interest.

### Funding Statement

This study was funded by UNFPA Morocco.

### Author Declarations

The study protocol was approved by the Ethics Committee of the Mohammed VI University of Sciences and Health (CE/UM6SS/63/24).

